# Examining the role of women’s engagement in khat production on child nutritional outcomes using longitudinal data in East Oromia, Ethiopia

**DOI:** 10.1101/2024.04.05.24305073

**Authors:** Karah Mechlowitz, Nitya Singh, Xiaolong Li, Dehao Chen, Yang Yang, Ibsa Abdusemed Ahmed, Jafer Kedir Amin, Abdulmuen Mohammed Ibrahim, Abadir Jemal Seran, Ibsa Aliyi Usmane, Arie H. Havelaar, Sarah L. McKune, the CAGED Research Team

**Affiliations:** Department of Family, Health and Wellbeing, University of Minnesota Extension, St. Paul, MN, United States of America; Department of Animal Sciences, Institute of Food and Agricultural Sciences, Emerging Pathogens Institute, Food Systems Institute, University of Florida, Gainesville, FL, United States of America; Emerging Pathogens Institute, Department of Environmental and Global Health, College of Public Health and Health Professions, University of Florida, Gainesville, FL, United States of America; Department of Epidemiology, Rollins School of Public Health, Emory University, Atlanta, GA, United States of America; Department of Statistics, Franklin College of Arts and Sciences, University of Georgia, Athens, GA, United States of America; Department of Public Health and Health Policy, School of Public Health, College of Health and Medical Sciences, Haramaya University, Haramaya, Oromia Regional State, Ethiopia; College of Veterinary Medicine, Haramaya University, Haramaya, Oromia Regional State, Ethiopia; Office of Research Affairs, Haramaya University, Haramaya, Oromia Regional State, Ethiopia; Haramaya University, Haramaya, Oromia Regional State, Ethiopia; School of Rural Development and Agricultural Innovation, College of Agriculture and Environmental Science, Haramaya University, Haramaya, Oromia Regional State, Ethiopia; Emerging Pathogens Institute, Global Food Systems Institute, Department of Animal Sciences, Institute of Food and Agricultural Sciences, University of Florida, Gainesville, FL, United States of America; Department of Environmental and Global Health, College of Public Health and Health Professions, Center for African Studies, University of Florida, Gainesville, FL, United States of America

## Abstract

In eastern Ethiopia, production of khat has increased in recent years, with significant implications for women in khat production and sale. Women have long been engaged in agricultural production in the region, yet the implications of the shift from food crop production to khat/cash crop production on degree and manner of women’s engagement in agriculture and any related changes in nutrition are largely unstudied. Using longitudinal data collected from December 2020 to June 2022 in Haramaya woreda, East Hararghe Zone, Ethiopia, this study aimed to explore the relationship between women’s engagement in khat production and child nutritional outcomes, and to test whether an increase/decrease in women’s engagement in khat production over time was associated with a change in child nutritional outcomes. Primary outcome variables were child length-for-age z-score (LAZ), child weight-for-age z-score (WAZ), child weight-for-length z-score (WLZ), and child minimum dietary diversity (MDD). Mixed effect models using backward stepwise regression were conducted to assess the relationship between women’s engagement in khat production, child nutrition outcomes, and a set of covariates, including women’s empowerment. No relationship was found between women’s engagement in khat production at baseline and child LAZ, WAZ, or WLZ; however, women’s empowerment was positively associated with child LAZ (β = 0.513, p = 0.004) and child WAZ (β = 0.456, p = 0.010) in this model. Women’s high engagement in khat production over time (first year of child’s life) was negatively associated with child LAZ (β = −0.731, p = 0.033) when compared to those who had low engagement over the same time period; women’s empowerment was positively associated with child LAZ (β = 0.693, p < 0.001) in this model. The findings from this study point to a potentially important dynamic between sustained high levels of women’s engagement in khat production and poor child growth outcomes; and contribute to a growing literature linking women’s empowerment in agriculture to improved child growth outcomes. Further research is needed to better understand the relationship between women’s engagement in khat production and women’s empowerment in agriculture.

## Introduction

The potential for agriculture to improve child nutritional outcomes in low- and middle-income countries (LMIC) is well established [1–4], as is evidence that women’s engagement in agriculture in Sub Saharan Africa (SSA) is important for food security, particularly within their households [5,6]. Best estimates indicate that nearly half of the economically active women in the world, and 79% in developing countries, report their primary activity as agriculture [7]. Recent efforts to make agricultural programming nutrition sensitive have demonstrated that through increased production, income, and women’s empowerment, agriculture has an important potential to improve nutritional outcomes [3,8]. However, there is also a significant body of evidence indicating that increasing income from agriculture alone is not necessarily sufficient to improve child nutritional outcomes [9,10]. When and where women make decisions about income allocation or agricultural production, child health and nutrition may improve; but increased investment in agriculture and improvement in agriculture-specific outcomes alone does not necessarily lead to improved child nutrition. Thus, it appears that where women are actively engaged in agricultural decision-making, children often have better nutritional outcomes [11–13].

In eastern Ethiopia, production of khat, a plant whose leaves are chewed as a recreational stimulant and in many cultural ceremonies, has increased exponentially in recent years and now dominates agricultural production in the Eastern Hararghe Zone, Oromia region. Women have long been engaged in agricultural production in the region, yet the implications of the shift from food crop production to khat/cash crop production on women’s engagement in agriculture and any consequent changes in food security and nutrition are largely unstudied. Previous research in the region found no relation between household engagement in khat production and child nutrition outcomes, but the study did not assess *women’s* engagement in khat production [14]. It did find associations between some domains of women’s empowerment in agriculture and child nutritional outcomes. Though women’s *empowerment* in agriculture is not the same as women’s *engagement* in agriculture, this finding underscores the question of whether women’s engagement in khat production in eastern Ethiopia might be influencing child nutritional outcomes. Thus, this paper seeks to explore the potential relationship between women’s engagement in khat production and child nutrition. By including women’s empowerment in the exploration, we will be able to better understand patterns of women’s roles, responsibilities, and agency in the context of differing livelihoods, particularly cash crop production and selling, thus improving our understanding of how agriculture to nutrition pathways may operate in a specific context.

The study is laid out as follows: a background section presents literature describing the contribution of women in agriculture and the agriculture-nutrition pathways of production, income, and women’s empowerment. Following, a short background on khat production in Ethiopia is presented, followed by methods, findings, and discussion.

## Background

### Women’s contribution to agriculture

From 1980 to 2010, women’s share of the population economically active in agriculture increased in SSA, as well as across all LMICs, providing an increasing evidence base for the “feminization of agriculture” hypothesis [15]. In SSA, women may grow a substantial share of staple crops in kitchen gardens and/or household plots, which are often not included in aggregate data on agriculture, despite these gardens (and labor needed to tend to them) playing an important role in household dietary diversity [5]. Women’s involvement in agriculture can vary depending on the specific crop and activity as well as the region. However, women tend to play a less active role in production of cash crops, where men’s presence dominates, thus limiting women’s engagement in income generating agricultural activities [16–18]. Evidence also indicates there may be shift in control from women to men when crops/commodities traditionally under women’s control are commercialized [16,17,19]. While women can still be involved in cash cropping, they may be involved to a lesser extent than men [16]. The commercialization of crops in smallholder farming households may have subtle but important implications on dimensions of women’s empowerment, particularly as livelihood strategies in LMIC evolve from subsistence-oriented to semi-subsistent or even commercially oriented agricultural landscapes [20,21].

In Ethiopia, gendered division of labor in cash cropping varies widely across locations and crop types. If timeliness is important, the workload to produce a cash crop may be shared between men and women; however, access to the benefits generated from production varies [22]. Additionally, women may be more involved in cash cropping and sales if fields and markets are closer to home [22].

### Agriculture to improved nutrition

In LMIC, livelihoods linked to agriculture contribute to the nutrition of household members through multiple, increasingly interactive pathways: food production, income, and women’s empowerment [23]. Household food production contributes to household nutrition by providing food for consumption and is the most direct pathway by which increased agricultural production translates to increased food availability and food security in smallholder farming households [24]. The type of food produced can influence diet quantity and quality, particularly for children, who have the highest risk of malnutrition. If a household owns livestock, products from livestock can also provide animal-source foods (ASFs), a source of high-quality nutrition critical for child growth and development [25]. The availability of land, labor, and technology drive the production level of different crops, which can affect patterns of household food consumption [24].

The second pathway, income from the sale of agricultural products, can assume a primary role in the agriculture to nutrition pathway when the market orientation of smallholder households increases [26]. Households may also engage in cash cropping to diversify income sources [27]. When the income pathway is more dominant, the type of food produced may depend on the household’s ability to sell it [10]. Income from agriculture sales can be used to purchase high quality foods, such as ASFs, and for other household necessities. Market access and prices and seasonality of crops can also change how households use production for consumption versus for sale [2,28,29]. Generally, though, smallholder farming households do not produce all the food a household needs, so production for consumption usually takes place alongside production for sale [28]. Most low-income smallholder households are net purchasers of food and may consume some staple foods that they produce while depending on local markets for other products [24]. Cash crops also play a different role in the agriculture to nutrition nexus, as they specifically contribute to the income pathway. While this can be a boom for farming communities, cash crop production can also push out food staple crops, reducing household food consumption [30].

Given the salience of the third agriculture-nutrition pathway—women’s empowerment— to the interpretation of results from this study, it is explored in more depth in the following section.

### Women’s empowerment and child nutrition

The third pathway, women’s empowerment, encompasses multiple dimensions, such as women’s agency and power, decision-making related to agriculture and income, time, labor, assets, knowledge, relationships, and intrahousehold food allocation [23]. There is a breadth of research investigating women’s empowerment and child nutrition, with many studies demonstrating positive relationships between women’s empowerment (or indicators of empowerment) and various child nutrition outcomes [9]. Studies in Ethiopia have demonstrated positive relationships between domains of women’s empowerment and child nutrition, including a positive association between empowerment in agricultural household decisions and increasing women’s access to and control of economic resources and stunting reduction [31]; a positive association between the five domains of empowerment (as measured by the Women’s Empowerment in Agriculture Index [WEAI]) and children’s dietary diversity [32]; a positive association between survey-based women’s empowerment index (SWPER) social autonomy and decision-making scores and the odds of meeting children’s minimum dietary diversity [33]; a positive association between disempowerment in decisions about input into production and child stunting [34]; and an association between enhanced empowerment in socioeconomic status (i.e. increasing access to education, information, media, and promoting saving) and the likelihood of child stunting or wasting [35]. There is growing evidence that targeting and empowering women through agriculture and livestock production may improve child nutrition [9]. When women are empowered, they are more equipped to positively impact their own health as well as their children’s [9,36,37].

### Conceptualizing and measuring women’s empowerment

The conceptualization of women’s empowerment, an oft-cited concept in gender and development work, differs across scholars and fields of study. The literature on empowerment is extensive and reflects evolving definitions, interpretations, and debates [38–47]. Scholars and practitioners have devised multiple approaches to measure women’s empowerment, including the Women’s Empowerment in Agriculture Index (WEAI), which was developed to quantitatively assess empowerment and gender inequality and compare across contexts to better understand connections between women’s empowerment, nutrition outcomes, and agriculture [48]. The WEAI measures five domains of women’s empowerment (production, resources, income, leadership, and time) through ten indicators. The WEAI has been widely used in agriculture and development work to measure the impacts of agricultural interventions on defined dimensions of women’s empowerment across contexts and time. Other indices have evolved from the WEAI that further explore empowerment, including the abbreviated WEAI (A-WEAI), the project WEAI (pro-WEAI), and Women’s Empowerment in Livestock Index (WELI) [49–53].

This current study will include women’s empowerment (as defined by the five domains of empowerment within the A-WEAI) in its analysis of the impact of women’s engagement in cash crop production on child nutritional outcomes. Although measuring empowerment using a tool such as the WEAI may not be inclusive of locally situated ideas and processes of empowerment or power relations between individuals and groups, it may provide insight into how women are involved in khat production and allow for better interpretation of any associations between women’s involvement and nutritional outcomes. For example, informal field discussions with researchers in the area indicated women may have more income under their control, yet spend more time away from weaning infants, if and when they are actively involved in the khat trade. Given the domains of women’s empowerment explored in the WEAI (production, resources, income, leadership, and time), the inclusion of the WEAI in models seeking to understand an association between women’s engagement and an improved or diminished nutritional outcome would be meaningful.

### Khat production in Ethiopia and nutrition

In eastern Oromia, Ethiopia, khat production and consumption are widespread and have been the subject of much debate [54]. The practice of chewing is culturally embedded and production and sale of khat as a cash crop drive local and regional economies [20,55]. Khat production in Harari and Oromia regions increased by 140% and 306%, respectively, between 2003 and 2017 [56]. Income from khat among smallholder farmers in Ethiopia has been estimated to be higher than that from many major agricultural crops, including income from grains/cereals, pulses, oilseeds, and coffee [57]. Based on a 2008-2009 agricultural survey, an estimated 2 million Ethiopian farmers cultivate khat, a number that has likely increased [57].

Khat chewing is widespread in the area and, though not the subject of this research, has been associated with undernutrition, restrictive dietary behaviors, and anemia in pregnant and lactating women [58,59]. A recent study also found that although parental khat use was a risk factor for poor child nutritional outcomes of stunting and wasting, maternal use was more strongly predictive, and the effect of khat chewing persisted regardless of household khat production or income. After khat chewing was considered, no relationship between khat production and child nutritional status remained [60].

Very few farmers in the area do not grow khat, as high profitability and drought tolerance have driven its widespread expansion [61]. One study, conducted in 10 woredas from Oromia and one other region (the Southern Nation, Nationality and People’s Region) found khat producers are much more likely to consume khat than non-producers [62]. It also found khat chewers substituted land holdings, including crop farms, backyard garden areas, pastoral, and other plots for khat production [62]. Khat’s economic growth as a cash crop and the potential for households to grow and harvest it year-round by using irrigation technology have made it economically vital to smallholder farming households in the region, as it has become a regular source of cash income [63].

The implications of this shift to the income pathway in the context of local khat/cash crop production on child nutrition are unknown. Previous research in the region found no significant associations between household engagement in khat production and child nutrition outcomes [14,60] but found associations between some domains of women’s empowerment and child nutrition [14]. While previous qualitative research indicated women’s involvement in khat production and sale may be significant [64], and informal discussions with local researchers confirm this, little has been documented about women’s involvement in khat production and sale in Ethiopia and any implications for child nutrition.

Using longitudinal data collected between December 2020 and June 2022, the specific objectives of this study are twofold: 1) to explore the relationship between women’s engagement in khat production and child nutritional outcomes, and, by leveraging the longitudinal data set 2) to test whether an increase in women’s engagement in khat production over time (one year) is associated with a change in child nutritional outcomes.

## Materials and methods

This study analyzed data collected from the Campylobacter Genomics and Environmental Enteric Dysfunction (CAGED) longitudinal cohort study. The CAGED study was designed to assess the fecal-oral transmission network of *Campylobacter* bacteria in Haramaya woreda, East Hararghe zone, Oromia region, Ethiopia, and to quantify the role of livestock, humans, and other reservoirs in this transmission. The study aims, research questions, and detailed methodology are published elsewhere [65], though an overview is briefly described below. This study was approved by the Institutional Review Board of the University of Florida, the Institutional Health Research Ethics Committee of Haramaya University, and the Ethiopia National Research Ethics Review Committee.

### Design and sample

A Health and Demographic Surveillance Site (HDSS) was established in 12 kebeles (smallest unit of government structure) in Haramaya woreda by Haramaya University (HU). Ten of these 12 kebeles provided the source population for the study. Using a birth registry, newborns were randomly selected to participate, with the aim to include 12 infants per kebele in the first month after birth; 115 infants were successfully enrolled. Enrollment began in December 2020 and ended in June 2021, with approximately 20 infants included each month. Written informed consent was obtained from all participating households (husband and wife) using a form in the local language (Afaan Oromo).

### Data collection

Infants were followed from birth to one year of age. Anthropometric measurements were collected once at enrollment, every three months during the study, and once at endline (month 13 for most enrolled infants). Trained data collectors from HU collected data on tablets using the REDCap mobile app [66,67]. Household questionnaires were administered to mothers and fathers at baseline and endline visits and contained questions on demographics; livelihoods; water, sanitation, and hygiene (WaSH); infant health and nutrition; wealth; animal ownership, management, and disease; and women’s empowerment. A short questionnaire containing questions on infant health, vaccinations, breastfeeding practices, antibiotic use, and diets was administered monthly to mothers. Stool samples were collected every four weeks from infants and biannually from mothers, siblings, and livestock.

### Variables

#### Outcome variables

Primary outcome variables of interest in this paper were child length-for-age z-score (LAZ), child weight-for-age z-score (WAZ), child weight-for-length z-score (WLZ), and child minimum dietary diversity (MDD). Child LAZ is a measure of linear growth faltering and can be an indicator of chronic undernutrition. A LAZ < −2 is considered stunted. All non-missing measurements for child LAZ for each participant were used, excluding LAZ at baseline due to unreliability in the data. Child WAZ is a growth outcome that can indicate that a child may be stunted, wasted, or both. A WAZ < −2 is considered underweight. All non-missing measurements for child WAZ for each participant were used. Child WLZ is growth outcome that can indicate recent and severe weight loss. A WLZ < −2 is considered wasted. All non-missing measurements for child WLZ for each participant were used. Child LAZ, WAZ, and WLZ were generated using validated methods for calculating anthropometric Z scores (using *anthro* package) [68,69]. Child MDD is an indicator designed by the World Health Organization to assess dietary diversity for children 6-23 months old as part of infant and young child feeding (IYCF) practices. Using a questionnaire, respondents (ideally the child’s caregiver) indicate whether their child consumed any food over the previous 24 hours from eight food groups (breast milk; grains, roots, and tubers; legumes and nuts; dairy products; flesh foods; eggs; vitamin A rich fruits and vegetables; other fruits and vegetables). Child MDD was calculated following published literature [70].

#### Women’s engagement in khat production

The primary explanatory variable for the first objective was women’s engagement in khat production as indicated at the baseline of the study. This was derived from women’s engagement score in khat production, which the research team created using six questions asked to women about their involvement in khat production. These six questions were recoded to produce a reduced set of four new questions describing women’s engagement in khat (Table 1). In the reduced set of questions, each of four recoded variables (involvement and ownership; frequency of market sale; use of income; decision-making about income) had possible values between 0 and 2, with value of two indicating “higher” engagement for that specific variable. An aggregated engagement score was then calculated by summing the values for each of the four engagement variables for each participant to indicate the extent of women’s engagement in khat production, with possible values ranging from 0 to 8. To strengthen the robustness of analysis, this score was then binarized based on the median engagement score to indicate higher engagement or no/low engagement in khat production for inclusion into the full model.

**Table 1.**
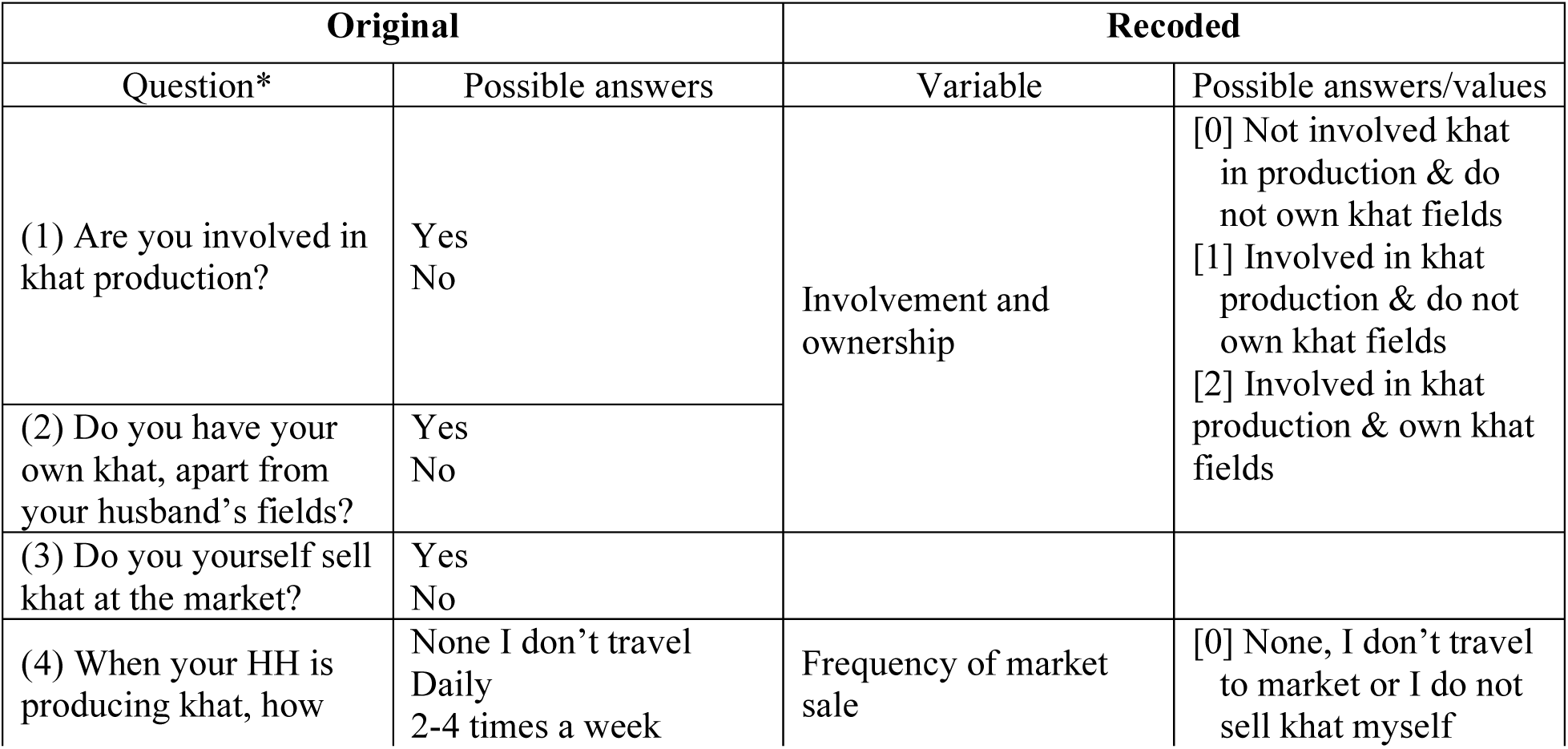
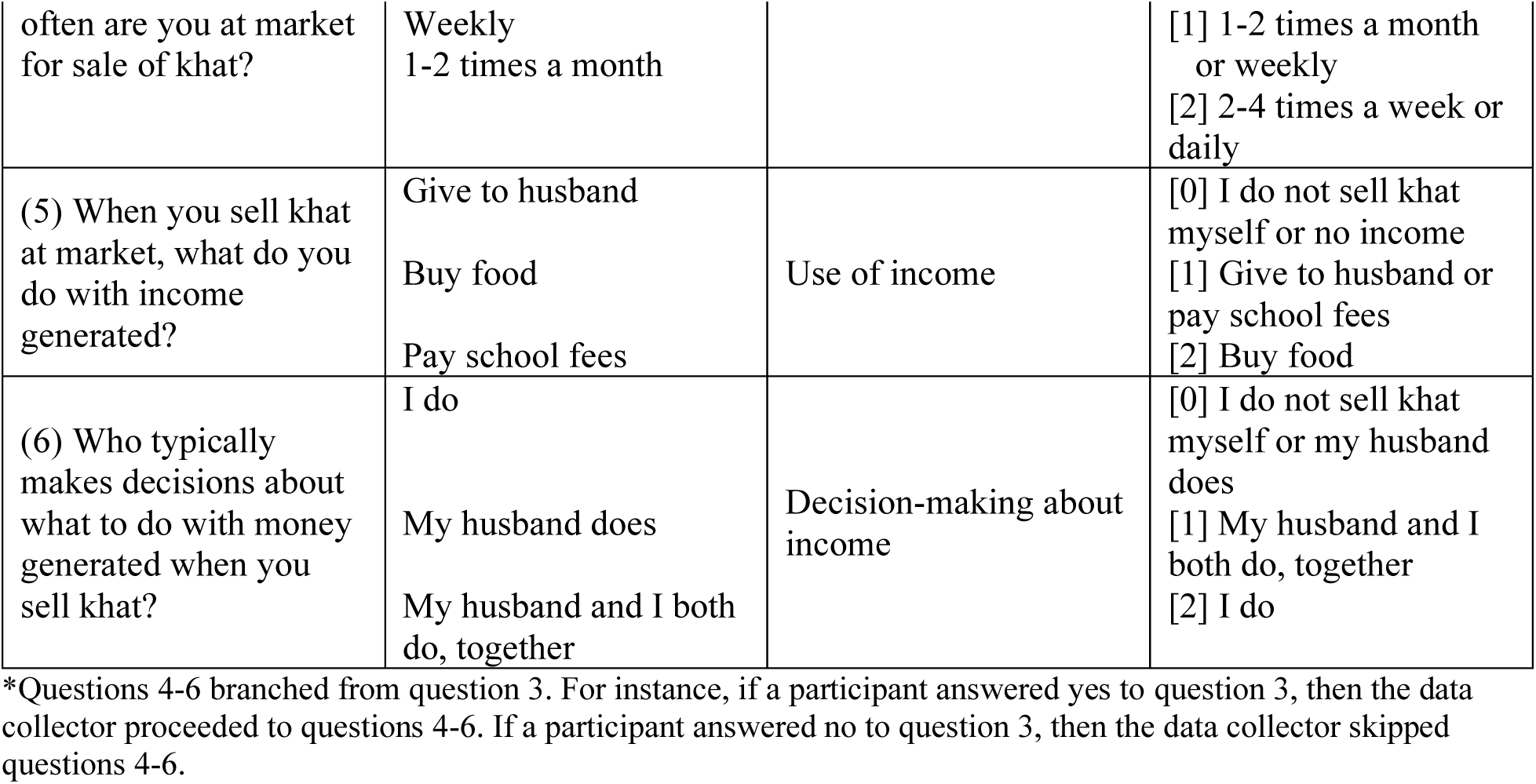
Original and recoded engagement in khat value chain questions used to generate engagement score.

#### Women’s engagement in khat production over time

The primary explanatory variable for the second objective was women’s engagement in khat production over time (from baseline to endline). Using the binarized engagement score noted above, we created four categories describing the change of consistency women’s engagement in khat production from baseline to endline (Table 2).

**Table 2.** Women’s khat production over time: categories of change in women’s engagement in khat production*.

| Women's engagement in khat production over time | Baseline engagement | Endline engagement |
| --- | --- | --- |
| Low engagement over time | No/low engagement | No/low engagement |
| Decreasing engagement | Higher engagement | No/low engagement |
| Increasing engagement | No/low engagement | Higher engagement |
| High engagement over time | Higher engagement | Higher engagement |

\*Derived from binarized women's engagement score in khat production

#### Covariates and confounders

The potential covariates selected for inclusion in the analysis included a household khat production ladder, mother’s age at baseline, women’s empowerment at baseline, tropical livestock unit (TLU) at baseline, exclusive breastfeeding (EBF) duration, mother’s school attendance, antenatal care while pregnant, household sanitation at baseline, and household hygiene at baseline. Confounders included household assets (quartiles) at baseline and child age and sex. The covariates and confounders initially selected were determined by existing literature and formative research in the study area [63,71].

A khat production ladder variable was constructed to represent household production intensity in khat production. The survey questions used to construct this variable included khat produced for livelihood, khat production as a primary means of livelihood, and khat production supported by irrigation in the off-season. Households could score between zero and three, with zero indicating no production in any of the above khat production activities. A score of one, two, or three indicated production intensity in one, two, or all three khat production activities, respectively.

Women’s empowerment, as measured by the five domains of empowerment (5DE) from the A-WEAI, was used in this analysis. Data on women’s empowerment were collected using the A-WEAI, which uses a validated questionnaire to assess women’s empowerment and inclusion in agriculture [72]. The A-WEAI contains two sub-indexes: the 5DE and the gender parity index (GPI). The 5DE measures the degree to which women are empowered in five domains: production, resource, income, leadership, and time. The 5DE is constructed from individual-level empowerment scores, which reflect each individual’s achievements in the five domains as measured by six indicators (productive decisions, ownership of assets, access to and decisions on credit, control over use of income, group membership, and workload) with their relative weights. The indicators assess if an individual has reached (or surpassed) adequate achievement with respect to each indicator, as defined by A-WEAI validated methods. Achieving adequacy in empowerment is defined by reaching adequacy in 80% or more of the weighted indicators. The GPI measures a woman’s achievements in the 5DE relative to the male counterpart in the household. The overall A-WEAI is a weighted average of the 5DE and GPI. The CAGED study collected the full A-WEAI at baseline and the 5DE for women at endline.

A TLU is a composite variable that factors livestock of different species by biomass in a household to allow for comparability among individuals. It was calculated following published literature [73]. Exclusive breastfeeding duration was defined as the last age (in days) that an infant, who was breastfed at birth, was reported to still be breastfeeding and to not have received anything other than breastmilk (prelacteal feed not considered in this definition; 78). Mother’s school attendance was treated as a binary variable (yes or no) defined as whether the mother ever attended school, given very low school attendance in the area. Antenatal care was binarized (yes or no) and defined as whether the mother ever received antenatal care while pregnant with her child.

Using the validated Joint Monitoring Program (JMP) ladder, sanitation ladder refers to the management of excreta from facilities used by people and includes the categories of safely managed, basic, limited, unimproved, and open defecation [75]. To strengthen robustness of analyses, household sanitation ladder was binarized; categories other than “open defecation” were combined. Using another validated JMP ladder, the hygiene ladder refers to the practices that contribute to maintaining health and preventing disease spread and includes the categories of basic, limited, and no facility [76]. To strengthen robustness of analyses, household hygiene ladder was binarized; categories other than “no facility” were combined. To calculate assets, following published literature [77–79], a latent trait model (using R package *ltm*) [80] was used to calculate a factor score for the binary responses indicating ownership of sellable household assets. The factor scores were then assigned to asset quartiles. To strengthen robustness of analyses, household asset quartile was binarized. The first and last two levels of household asset quartile were collapsed to indicate low or high household assets, respectively.

### Analysis

Descriptive analyses of sociodemographic variables were performed to explore characteristics of the study population and to characterize khat production at baseline and endline. Because of non-normality and small sample size, a Wilcoxon signed-rank test was performed to test for a significant difference in engagement score in khat production from baseline to endline. For each outcome, a linear mixed model with individual level random intercept was used to account for non-independence among repeated observations using the R package *lme4* [81].

Potential covariates were tested in unadjusted and adjusted bivariate analyses using a linear mixed model with an individual level random intercept to determine which covariates were entered into the full mixed effect model. Covariates were selected for inclusion into the full mixed effect models if the adjusted p-value was < 0.25 (adjusted for child age, infant sex, and binarized household assets). Engagement in khat production and change in engagement in khat production were included in their respective full mixed effect models given their central importance to the objectives of this paper. Backward stepwise regression was used to determine the final model for each objective (p-value < 0.05). Data analysis was performed in R statistical software version 4.2.2 [82].

## Results

A total of 106 infants were included in the analysis. Univariate summaries for various characteristics at baseline are shown in Table 3. Mean mother’s age was 27 years, and the mean TLU was 0.86. The average number of days an infant was exclusively breastfed was 60.6 days. Fifty-six percent of women were empowered, and 27.1% of mothers attended school. Regarding the change in women’s engagement in khat production from baseline to endline, 52.7% of women had low engagement over time, 12.1% had decreasing engagement, 26.4% had increasing engagement, and 8.8% had higher engagement over time.

**Table 3.** Univariate summaries for baseline household characteristics.

| Characteristic | Univariate Summary<sup>^</sup> |
| --- | --- |
| Mother's age* | 27.0 (10.4) |
| TLU*<sup>&</sup> | 0.88 (1.2) |
| Exclusive breastfeeding duration (days) | 60.6 (89) |
| Women's empowerment |  |
| Empowered | 56.5 (52) |
| Not empowered | 43.5 (40) |
| Infant being female* | 48.1 (51) |
| Mother attending school* | 27.1 (26) |
| JMP Sanitation ladder*<sup>&</sup> |  |
| Open defecation | 70.4 (69) |
| Unimproved | 18.4 (18) |
| Limited | 2.0 (2) |
| Basic | 9.2 (9) |
| JMP Hygiene ladder*<sup>&</sup> |  |
| No facility | 74.2 (72) |
| Limited | 5.2 (5) |
| Basic | 20.6 (20) |

\*Data previously published in (Chen et al., under review) but are presented here to provide a demographic understanding of the study population. <sup>^</sup>Percentage (frequency) presented for categorical variables. Mean (interquartile range) presented for continuous variables. <sup>&</sup>TLU: tropical livestock units; JMP: WHO/UNICEF Joint Monitoring Program for Sanitation and Hygiene.

Descriptive statistics for khat production are shown in Table 4. At baseline and endline, the majority of households reported engaging in khat production as a livelihood activity. At baseline, 14.7% of women reported being involved in khat production and not owning khat fields, while 81.2% of women reported being involved in khat production and not owning khat fields at endline. At endline, more women were traveling to the market to sell khat (34.7%) thanat baseline (12.6%). The Wilcoxon signed-rank test indicated there is a significant difference in women’s engagement score in khat production over time (from baseline and endline (p < 0.05).

**Table 4.**
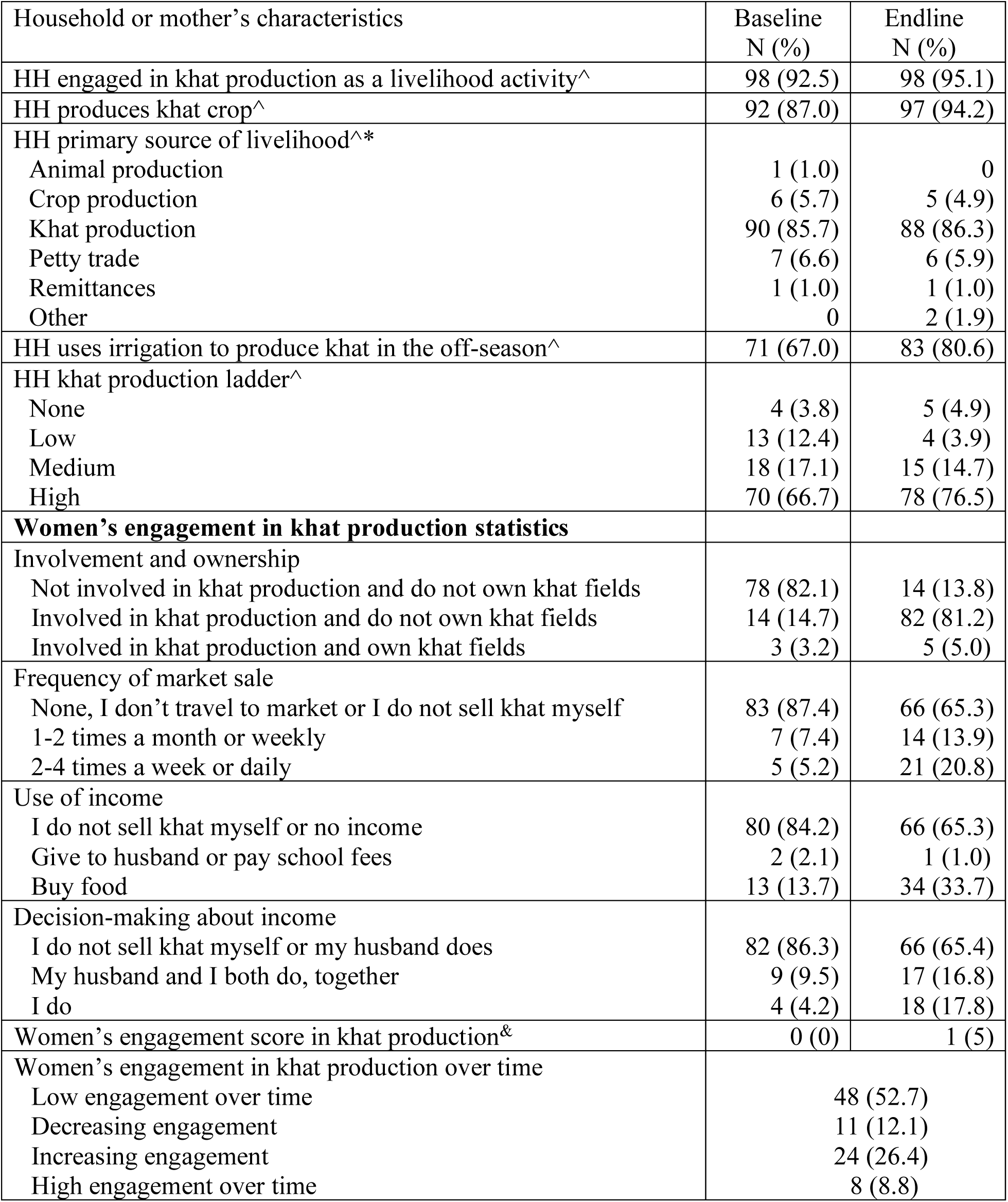
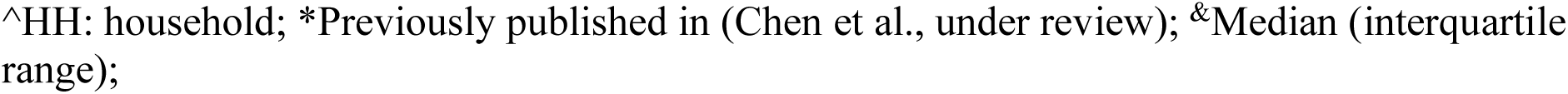
Descriptive statistics for khat production at baseline and endline.

### Unadjusted and adjusted mixed-effect model results

Table 5 displays the results for the unadjusted and adjusted mixed effect models for each covariate and the outcomes of LAZ, WAZ, WLZ, and MDD. For the outcome of LAZ, in the adjusted model (Table 5), women’s empowerment was significantly positively associated with LAZ (β = 0.513, p = 0.04). Women’s empowerment, TLU, and sanitation ladder met the inclusion criteria for the full linear mixed models. For the outcome of WAZ, in the adjusted model (Table 5), women’s empowerment was significantly positively associated with WAZ (β = 0.456, p = 0.010). Tropical livestock unit was significantly positively associated with WAZ (β = 0.207, p = 0.022). Women’s empowerment, TLU, and sanitation ladder met the inclusion criteria for the full linear mixed models.

**Table 5.**
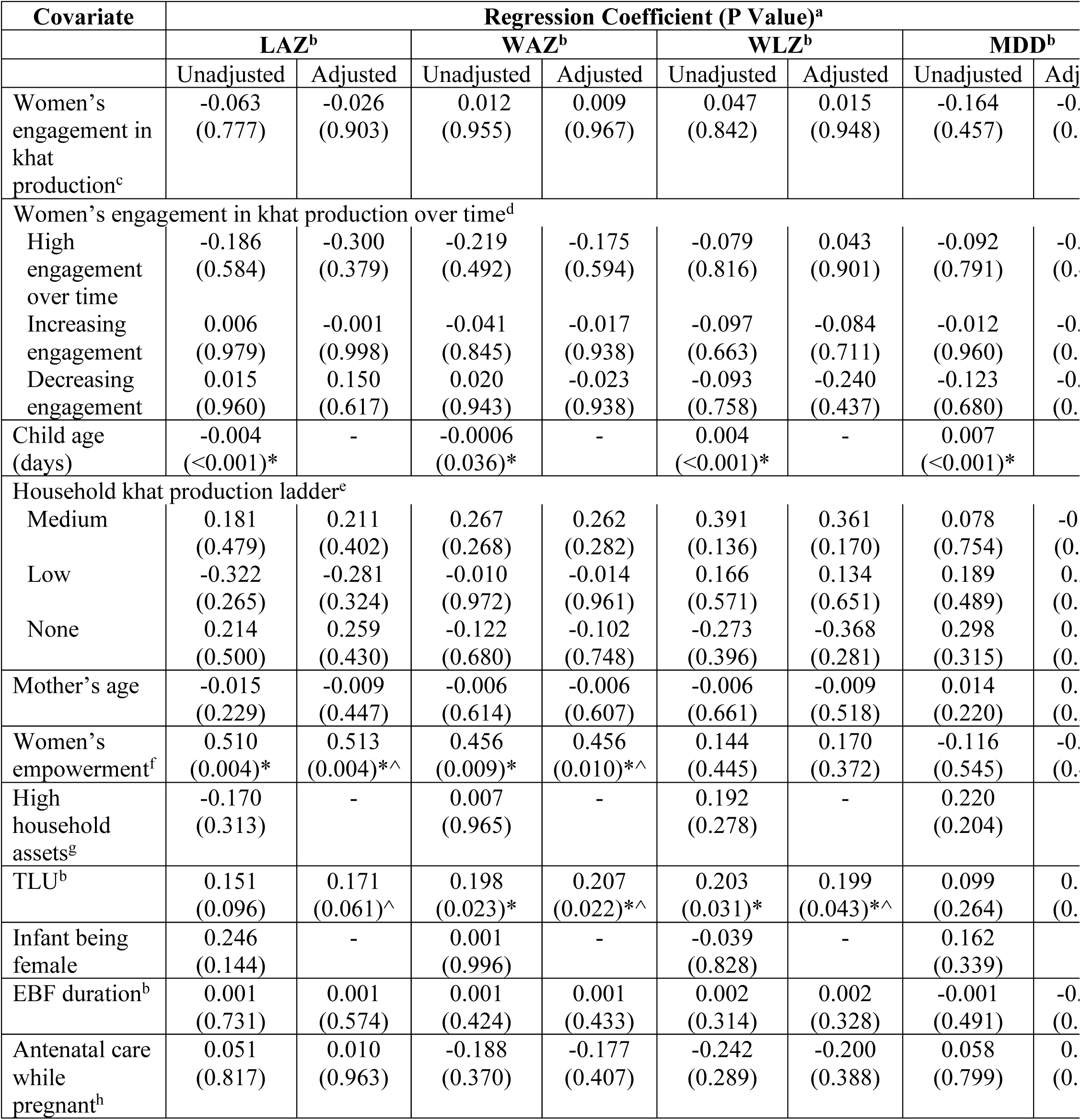
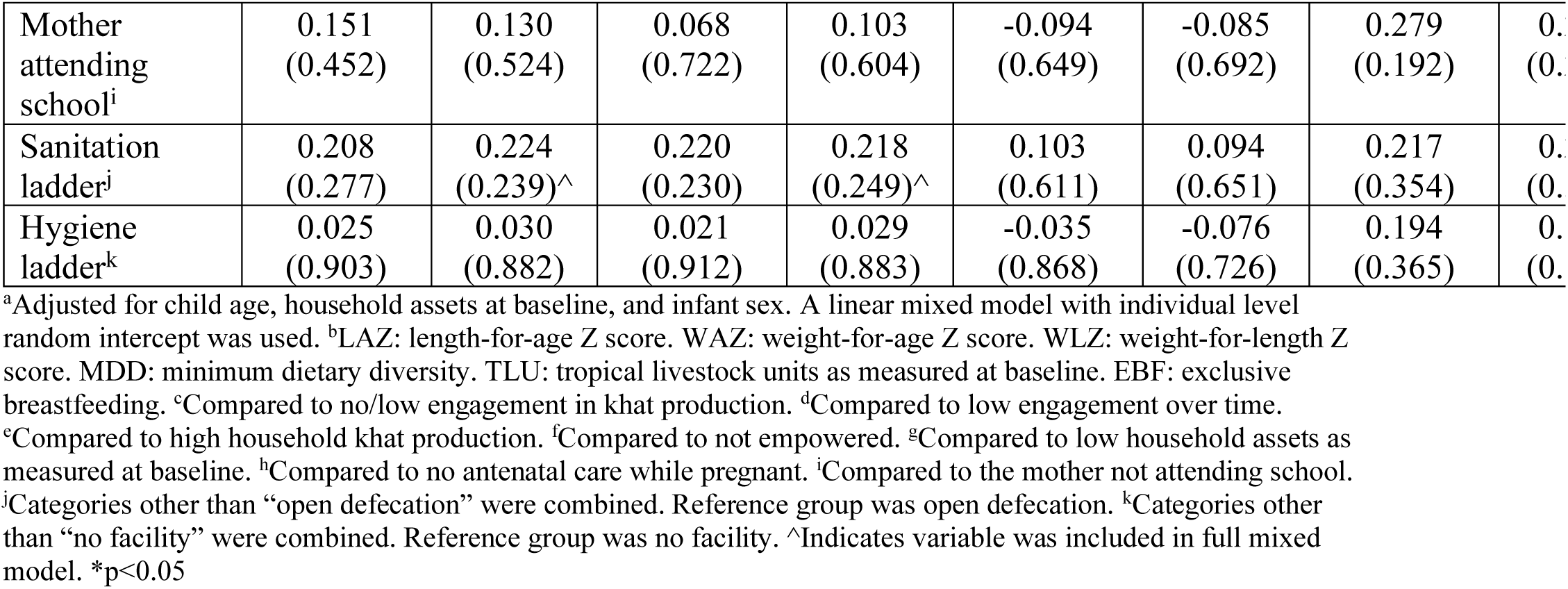
Unadjusted and adjusted univariable mixed effect model results for the child outcomes of length-for-age Z score, weight-for-age Z score, weight-for-length Z score, and minimum dietary diversity.

For the outcome of WLZ, in the adjusted analyses (Table 5), TLU was significantly positively associated with WLZ (β = 0.199, p = 0.043). Tropical livestock unit was the only variable that met the inclusion criteria for the full linear mixed model. For the outcome of MDD, in the bivariate analyses (Table 5), none of the covariates met the inclusion criteria for entry into the full linear mixed effect model.

### Full mixed-effect model analyses

#### Women’s engagement in khat production

In the full linear mixed model investigating women’s engagement in khat production at baseline and child LAZ (Table 6), after backward selection, women’s empowerment was positively associated with child LAZ (β = 0.513, p = 0.004).

**Table 6.**
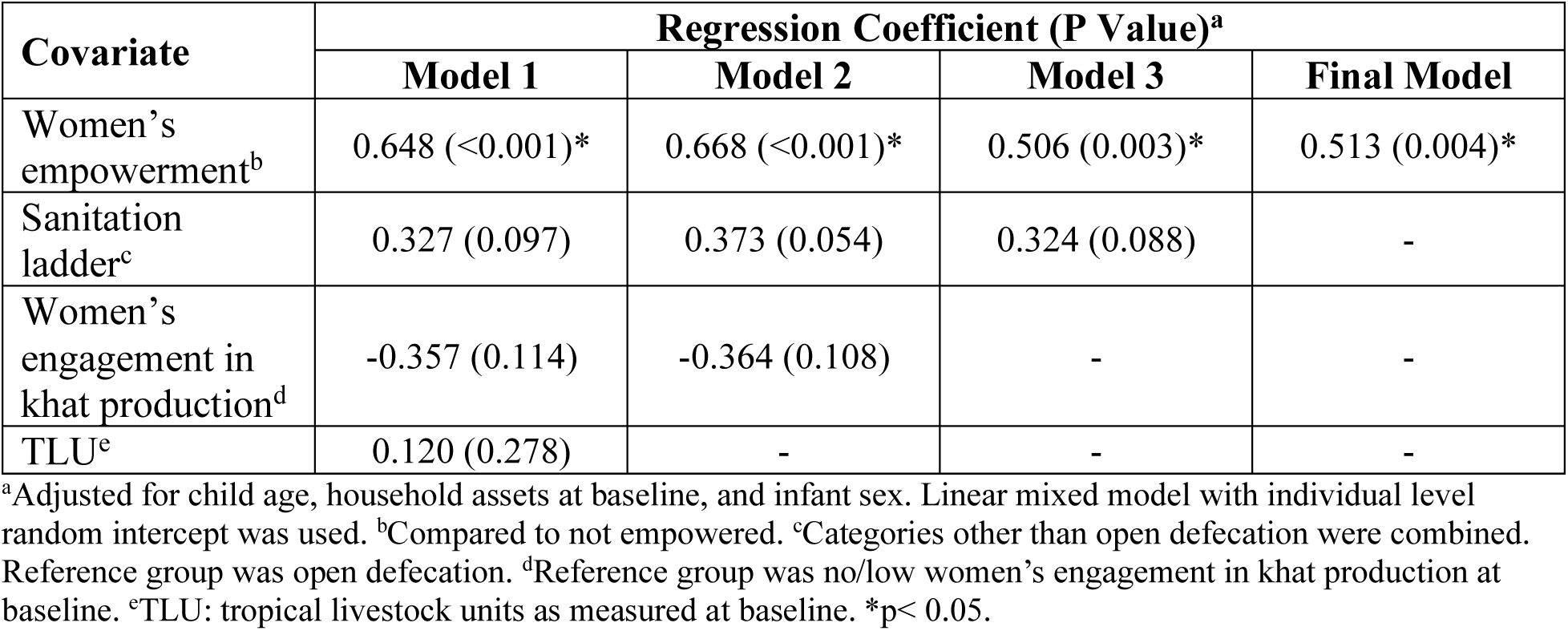
Backward stepwise regression results between women’s engagement in khat production, covariates, and the outcome of child length-for-age Z score.

| Covariate | Regression Coefficient (P Value)<sup>a</sup> |  |  |  |
| --- | --- | --- | --- | --- |
|  | Model 1 | Model 2 | Model 3 | Final Model |
| Women's empowerment<sup>b</sup> | 0.648 (<0.001)* | 0.668 (<0.001)* | 0.506 (0.003)* | 0.513 (0.004)* |
| Sanitation ladder<sup>c</sup> | 0.327 (0.097) | 0.373 (0.054) | 0.324 (0.088) | - |
| Women's engagement in khat production<sup>d</sup> | -0.357 (0.114) | -0.364 (0.108) | - | - |
| TLU<sup>e</sup> | 0.120 (0.278) | - | - | - |

<sup>a</sup>Adjusted for child age, household assets at baseline, and infant sex. Linear mixed model with individual level random intercept was used. <sup>b</sup>Compared to not empowered. <sup>c</sup>Categories other than open defecation were combined. Reference group was open defecation. <sup>d</sup>Reference group was no/low women's engagement in khat production at baseline. <sup>e</sup>TLU: tropical livestock units as measured at baseline. \* $p < 0.05$ .

In the full linear mixed model investigating women’s engagement in khat production at baseline and child WAZ (Table 7), after backward selection, women’s empowerment remained positively associated with child WAZ (β = 0.456, p = 0.010), while TLU was no longer significant.

**Table 7.** Backward stepwise regression results between women’s engagement in khat production, covariates, and the outcome of child weight-for-age Z score.

| Covariate | Regression Coefficient (P Value)<sup>a</sup> |  |  |  |
| --- | --- | --- | --- | --- |
|  | Model 1 | Model 2 | Model 3 | Final Model |
| Women's empowerment<sup>b</sup> | 0.515 (0.009)* | 0.430 (0.014)* | 0.449 (0.010)* | 0.456 (0.010)* |
| Sanitation ladder<sup>c</sup> | 0.274 (0.178) | 0.281 (0.149) | 0.325 (0.090) | - |
| TLU<sup>d</sup> | 0.142 (0.216) | 0.132 (0.233) | - | - |
| Women's engagement in khat production<sup>e</sup> | -0.194 (0.404) | - | - | - |

<sup>a</sup>Adjusted for child age, household assets at baseline, and infant sex. Linear mixed model with individual level random intercept was used. <sup>b</sup>Compared to not empowered. <sup>c</sup>Categories other than open defecation were combined. Reference group was open defecation. <sup>d</sup>TLU: tropical livestock units as measured at baseline. <sup>e</sup>Reference group was no/low women's engagement in khat production. \*p< 0.05

In the full linear mixed model investigating women’s engagement in khat production at baseline and child WLZ (Table 8), after backward selection, only TLU remained positively associated with child WLZ (β = 0.199, p = 0.043).

**Table 8.**
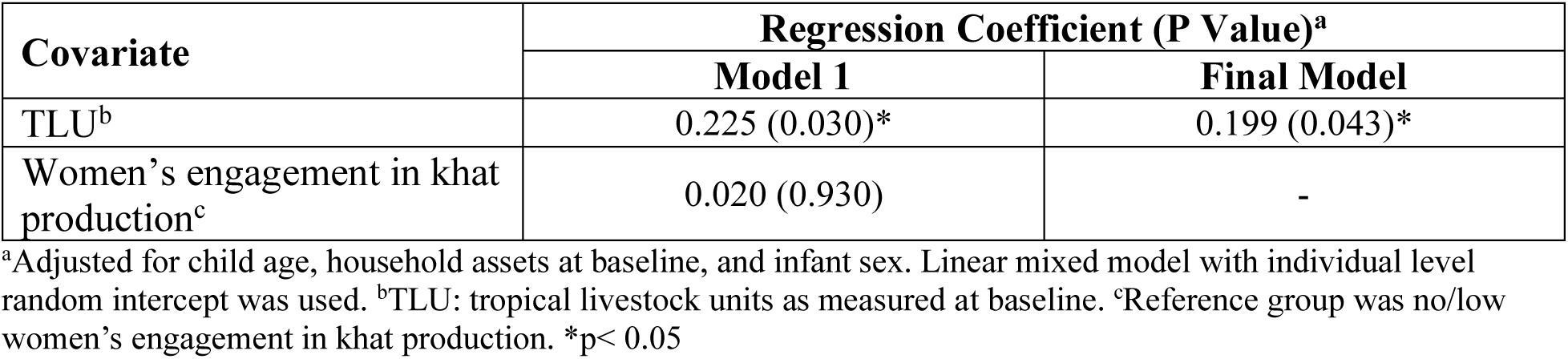
Backward stepwise regression results between women’s engagement in khat production, covariates, and the outcome of child weight-for-length Z score.

| Covariate | Regression Coefficient (P Value)<sup>a</sup> |  |
| --- | --- | --- |
|  | Model 1 | Final Model |
| TLU<sup>b</sup> | 0.225 (0.030)* | 0.199 (0.043)* |
| Women's engagement in khat production<sup>c</sup> | 0.020 (0.930) | - |

<sup>a</sup>Adjusted for child age, household assets at baseline, and infant sex. Linear mixed model with individual level random intercept was used. <sup>b</sup>TLU: tropical livestock units as measured at baseline. <sup>c</sup>Reference group was no/low women's engagement in khat production. \*p< 0.05

#### Women’s engagement in khat production over time

In the full linear mixed model investigating women’s engagement in khat production over time and child LAZ (Table 9), after backward selection, women with high engagement in khat production over time was associated with a decrease in child LAZ when compared to those who had low engagement over time (β = −0.73, p = 0.033). Additionally, women’s empowerment was associated with an increase in child LAZ (β = 0.693, p = 0.001).

**Table 9.** Backward stepwise regression results between women’s engagement in khat production over time, covariates, and the outcome of child length-for-age Z score.

| Covariate | Regression Coefficient (P Value)<sup>a</sup> |  |  |
| --- | --- | --- | --- |
|  | Model 1 | Model 2 | Final Model |
| Women's empowerment<sup>b</sup> | 0.663 (0.001)* | 0.679 (<0.001)* | 0.693 (<0.001)* |
| Women's engagement in khat production over time<sup>c</sup> |  |  |  |
| High engagement | -0.733 (0.031)* | -0.757 (0.025)* | -0.731 (0.033)* |
| Increasing engagement | -0.101 (0.633) | -0.103 (0.625) | -0.151 (0.476) |
| Decreasing engagement | -0.177 (0.557) | -0.169 (0.572) | -0.140 (0.646) |
| Sanitation ladder<sup>d</sup> | 0.344 (0.095) | 0.379 (0.061) | - |
| TLU<sup>e</sup> | 0.094 (0.408) | - | - |

<sup>a</sup>Adjusted for child age, household assets at baseline, and infant sex. Linear mixed model with individual level random intercept was used. <sup>b</sup>Compared to not empowered. <sup>c</sup>Reference group was low engagement over time.
<sup>d</sup>Categories other than open defecation were combined. Reference group was open defecation. <sup>e</sup>TLU: tropical livestock units as measured at baseline. \*p< 0.05.

In the full linear mixed model investigating women’s engagement in khat production over time and child WAZ (Table 10), after backward selection, only women’s empowerment remained positively associated with child WAZ (β = 0.456, p = 0.010), while TLU was no longer significant.

**Table 10.**
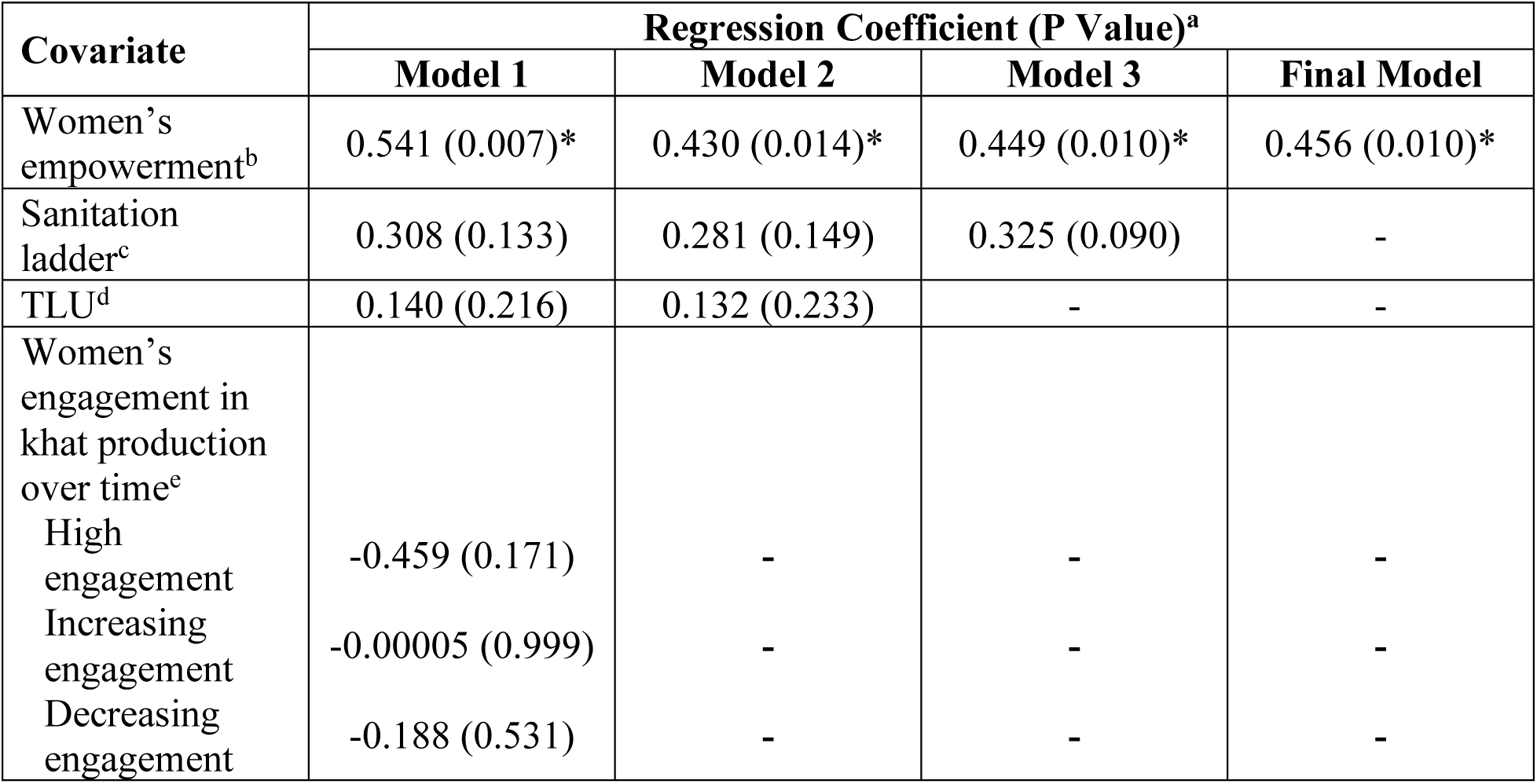
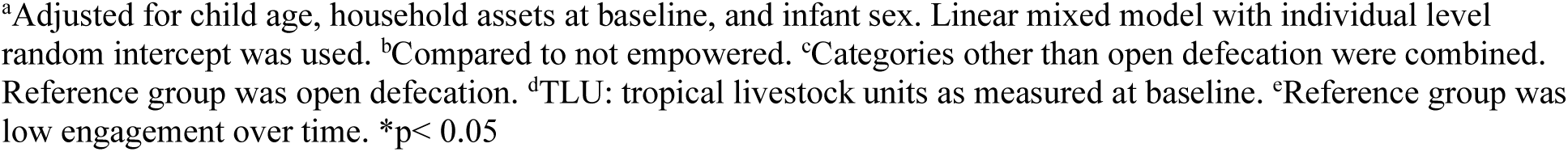
Backward stepwise regression results between women’s engagement in khat production over time, covariates, and the outcome of child weight-for-age Z score.

| Covariate | Regression Coefficient (P Value)<sup>a</sup> |  |  |  |
| --- | --- | --- | --- | --- |
|  | Model 1 | Model 2 | Model 3 | Final Model |
| Women's empowerment<sup>b</sup> | 0.541 (0.007)* | 0.430 (0.014)* | 0.449 (0.010)* | 0.456 (0.010)* |
| Sanitation ladder<sup>c</sup> | 0.308 (0.133) | 0.281 (0.149) | 0.325 (0.090) | - |
| TLU<sup>d</sup> | 0.140 (0.216) | 0.132 (0.233) | - | - |
| Women's engagement in khat production over time<sup>e</sup> |  |  |  |  |
| High engagement | -0.459 (0.171) | - | - | - |
| Increasing engagement | -0.00005 (0.999) | - | - | - |
| Decreasing engagement | -0.188 (0.531) | - | - | - |

<sup>a</sup>Adjusted for child age, household assets at baseline, and infant sex. Linear mixed model with individual level random intercept was used. <sup>b</sup>Compared to not empowered. <sup>c</sup>Categories other than open defecation were combined. Reference group was open defecation. <sup>d</sup>TLU: tropical livestock units as measured at baseline. <sup>e</sup>Reference group was low engagement over time. \*p< 0.05

In the full linear mixed model investigating the change in women’s engagement in khat production over time and child WLZ (Table 11), after backward selection, only TLU remained positively associated with WLZ (β = 0.199, p = 0.043).

**Table 11.** Backward stepwise regression results between women’s engagement in khat production over time, covariates, and the outcome of child weight-for-length Z score.

| Covariate | Regression Coefficient (P Value)<sup>a</sup> |  |
| --- | --- | --- |
|  | Model 1 | Final Model |
| TLU<sup>b</sup> | 0.257 (0.010)* | 0.199 (0.043)* |
| Women's engagement in khat production over time<sup>c</sup> |  |  |
| High engagement | 0.112 (0.738) | - |
| Increasing engagement | -0.042 (0.846) | - |
| Decreasing engagement | -0.259 (0.387) | - |

<sup>a</sup>Adjusted for child age, household assets at baseline, and infant sex. Linear mixed model with individual level random intercept was used. <sup>b</sup>TLU: tropical livestock units as measured at baseline. <sup>c</sup>Reference group was low engagement over time. \*p< 0.05.

## Discussion

This study utilized survey data to investigate how women’s engagement in khat production and sale is related to child nutrition outcomes in eastern Ethiopia and what, if any, role women’s empowerment plays. In recent years, khat production has been increasingly commercialized and is a dominant cash crop in the Oromia region in eastern Ethiopia [20,63]. Smallholder farmers in Haramaya, Ethiopia, often practice food-based crop production alongside khat production to generate produce and khat for food and cash [20,57,83]. The results from this study show that women’s engagement in khat production was much greater at endline compared to baseline. This difference in women’s involvement from baseline to endline may represent a real temporal increase in women’s engagement in khat or it may reflect cultural norms around women’s behavior immediately following childbirth. This is explored in more depth below.

Enrollment for this study began in December 2020, a time when COVID-19 restrictions and associated lockdowns could have limited women’s engagement in production and traveling to markets for selling khat. Importantly, though, political unrest and tension that began, in part, due to delayed national elections earlier in 2020 sparked a major conflict between Tigray People’s Liberation Front (TPLF), an ethnic nationalist group in Ethiopia, and the Ethiopian federal government, which ballooned into a two-year war that began in November 2020. The effects of this war reverberated throughout Ethiopia, and the resulting tensions could have contributed to less engagement in activities outside the home or village.

Complicating interpretation of these data, the baseline data were collected when women had just given birth, limiting their engagement in activities other than caring for their infant. Some research on postpartum practices in Ethiopia indicate there is often a period of seclusion, in which mothers may not leave the house for a period of time after giving birth [84,85]. The endline data may indicate a level of involvement in khat production that is more typical and less characterized by postpartum practices, COVID-19-related lockdowns, and political unrest and conflict. Furthermore, rising inflation, particularly for food prices, and slowing economic growth globally have made it challenging for smallholder farming households to afford food [86]. This dramatic increase in engaging in khat production occurred over a period of increasing food insecurity, and anecdotes support these findings that women began actively engaging in household khat production activities due to new exogenous stresses. This added pressure may have contributed to the need to diversify income sources and driven more women to khat production and sale.

While women’s engagement in khat production at baseline was not associated with LAZ, WAZ, or WLZ, women who had high engagement in khat production over time (meaning women who had higher engagement in khat production from baseline to endline) did have worse child LAZ outcomes than women who had low engagement in khat production over time. Furthermore, when looking closely at the relationship between higher engagement in khat production at baseline and child LAZ (Table 6), there is some indication of a potential, albeit non-significant, negative relationship between higher engagement in khat production at baseline and child LAZ (β = −0.364, p = 0.108). Women’s empowerment, however, was consistently significantly positively associated with LAZ and WAZ, meaning women who were empowered had better child LAZ and WAZ outcomes than women who were not empowered. Given these findings, additional analyses investigating the relationship between women’s engagement in khat production and women’s empowerment were conducted (S1 Table). Interestingly, these additional findings indicate no relationship between women’s engagement in khat production and women’s empowerment, although this analysis was limited by the small sample size.

One possible explanation for the relationship between women’s high engagement in khat production over time and child LAZ may relate to women’s time spent producing, traveling to the market, and selling khat, taking time away from directly caring for their children. This time trade-off could lead to reduced breastfeeding duration, suboptimal complementary feeding practices, and inadequate supervision of young children, all of which are crucial for ensuring optimal growth and development during the critical early years. This is particularly important given that the first 1000 days of life represent a critical window of human development, where poor nutrition and inadequate care can have short- and long-lasting implications on human health and function, including stunting [87]. These findings are also particularly relevant within the context of recent inflation throughout Ethiopia. Economic recessions and fluctuations in income often contribute to increased participation by women in agricultural labor, with potentially harmful effects on child survival [88,89]. This may be related, in part, to the tradeoff between women’s time in reproductive work (e.g., childcare and domestic tasks) vs. in agriculture.

Although many women are economically engaged in agricultural labor, they tend to maintain unequal childcare and domestic responsibilities relative to men and may have to conduct paid agricultural work alongside their unpaid domestic labor, as opposed to sequentially performing such duties [90,91]. Additionally, if women’s workload in agricultural labor is not matched by enhanced decision-making power and control of resources, then maternal and child nutrition may suffer [92]. This is particularly important to consider given khat’s role as a cash crop in eastern Oromia and the dynamics that occur between men and women in agricultural activities involving cash crops.

The results from this study suggest income (and other benefits) generated from engaging in khat production may not be enough to offset the possible detrimental effect of how much time women spend in khat production and sale. In some cases, income generated from khat sales may not be allocated towards improving household food security or investing in children’s nutrition and healthcare needs. Instead, it could be spent on other expenses or priorities, further exacerbating the nutritional vulnerability of children within these households. While these results point to a potentially negative relationship between women’s high engagement in khat production over time and child LAZ, they do not pinpoint what domains of engagement (e.g., income use, decision-making, and/or ownership) are most important for child nutrition. Indeed, gendered division of labor in agriculture is dynamic and can change in response to commercialization and innovation [93,94], and tradeoffs related to women’s time use are complex and unpredictable [89,90].

The state of the literature on women’s empowerment and child nutrition demonstrates that women’s empowerment is generally positively associated with child nutrition, but that it can be difficult to interpret and compare results due to various empowerment indicators and empowerment categorizations [95–98]. Furthermore, few studies investigating women’s empowerment and HAZ included children under two years of age [95]. Thus, the findings in this study represent an important contribution to understanding the relationship between women’s empowerment and infant growth outcomes. A 2019 systematic review investigating the role of women’s empowerment in child nutrition outcomes found that, of the 461 associations identified from 39 studies, 70 associations were statistically significantly positive, and 186 of 307 associations for HAZ were between −0.10 and 0.10 standard deviations [95]. A study conducted in northwest Ethiopia investigating women’s empowerment (as measured by five different empowerment indicators) and child stunting found that the odds of having stunted children were 44% lower among mothers with moderate empowerment compared to mothers with low empowerment [99]. Another study using WEAI data from six countries in Africa and Asia found that various WEAI indicators, including more agricultural decisions, a higher number of agricultural assets with rights, a higher number of credit decisions, and greater satisfaction with leisure along with the aggregated WEAI score were associated with higher child HAZ [100].

Interestingly, for the outcome of WLZ, only TLU was positively associated with child WLZ, meaning that a higher household TLU was related to better child WLZ outcomes. Like agriculture production, livestock production can be linked to improved child nutrition outcomes through the pathways of empowerment, food production, and income by providing nutrient-rich animal source foods (ASFs) and income generation that can be used to purchase food and provide access to better healthcare, education, and sanitation [9,101]. A WLZ < −2 is an indicator of wasting, a more acute condition resulting from recent rapid weight loss, thus making it more responsive to changes in diet and/or changes in treatment.

The literature on livestock ownership and child nutrition outcomes in LMIC is robust; however, studies investigating livestock ownership and child WLZ in LMIC have found mixed results. In Uganda, a weak association was found between livestock ownership and the probability of a child being wasted, although this effect was limited to children between two and five years of age [102]. Improved dairy cow adoption in Uganda, while found to be associated with child HAZ, was not associated with child WHZ [103]. Azzarri et al. (2015) found that livestock ownership, specifically small ruminants, was related to a lower probability of being wasted (defined as WLZ < −2) in Uganda [104]. No effect was found between co-owned/female-owned livestock and child WLZ in Kenya, while a positive association was found between co-owned/female-owned livestock and child WAZ with a mediating effect by child ASF intake [105]. Similarly, no relationship between livestock ownership and child WLZ was found in western Kenya [106]. Given the multi-level determinants of child anthropometric outcomes, a combination of efforts, including the promotion of optimal livestock keeping practices to maintain a healthy household-animal environment and gender-sensitive interventions aimed to increase child and household nutrition outcomes, are needed to translate livestock production into optimal consumption of ASF and improved child nutrition [9,107]. While this study contributes to the growing body of literature on women’s empowerment and child nutrition outcomes, further comparative analysis with similar studies is warranted to validate our findings and elucidate potential contextual differences. Additionally, exploring potential moderators or mediators of this relationship, such as cultural norms, social support networks, or access to healthcare services, could provide valuable insights into the mechanisms underlying these associations.

This paper has some limitations. The CAGED study was powered to different outcomes of interest; as such, the objective of this paper was not to detect significance but rather to explore patterns of women’s engagement in khat production, an economically and culturally important crop in the study region, to better understand how women’s involvement in an economically beneficial activity, and other important sociodemographic factors, may relate to child nutrition outcomes. Despite adjusting for confounders of child sex, child age, and household assets, there could be other potential confounders not accounted for by this survey-based study that could bias the findings in this paper. Lastly, the risk of social desirability can be higher in situations where the literacy rate is low (as in the CAGED study participants) and where there is limited freedom of speech [108]. To mitigate the risk of social desirability bias, all enumerators in the CAGED study were from the local community, aware of cultural norms and beliefs, and adopted various strategies aimed at building trust [109].

## Conclusion

In conclusion, this study builds on the CAGED formative research, offering a more comprehensive exploration and understanding of the complex dynamics between women’s engagement in khat production, rural livelihoods, and child nutrition outcomes, and reinforcing earlier findings that point to the importance of women’s empowerment for child nutrition outcomes. The findings point to a potentially important dynamic between high levels of women’s engagement in khat production over time and reduced child length-for-age z-scores. Additional research is needed to confirm this finding, yet it illustrates the complexity of any policy or programmatic intervention: how to encourage the benefits of women’s empowerment that may come from engagement in cash cropping, while concurrently mitigating the possible detrimental impacts on children in the home. These findings underscore the importance of assessing women’s time burden as an outcome of any agricultural policy or program aiming to improve nutritional outcomes and the importance of understanding the gendered patterns of labor in agriculture and household food security or nutrition. Future research should include a more nuanced, qualitative exploration of empowerment in agriculture using methodological tools that can capture how women integrate into khat and other cash crop value chains.

## Data Availability

Deidentified individual participant data will be made available through Dataverse (https://dataverse.org/) after December 31, 2024.

## Acknowledgements

This work is a result of the CAGED Research Team whose members include: Amanda Evelyn Ojeda, Arie H. Havelaar, Abadir Jemal Seran, Abdulmuen Mohammed Ibrahim, Bahar Mummed Hassen, Belisa Usmael Ahmedo, Cyrus Saleem, Dehao Chen, Efrah Ali Yusuf, Gireesh Rajashekara, Getnet Yimer, Ibsa A. Ahmed, Ibsa Aliyi Usmane, Jafer Kedir Amin, Jemal Y. Hassen, Kedir A. Hassen, Kunuza Adem Umer, Karah Mechlowitz, Kedir Teji Roba, Loic Deblais, Mussie Bhrane, Mark J. Manary, Mawardi M. Dawid, Mahammad Mahammad Usmail, Nigel P. French, Nur Shaikh, Nitya Singh, Sarah L. McKune, Wondwossen A. Gebreyes, Xiaolong Li, Yenenesh Demisie Weldesenbet, Yang Yang, and Zelalem Hailu Mekuria.

## Supporting information

**S1 Table. Chi-square or Fisher’s exact test results for women’s engagement in khat production and women’s empowerment.** ^Indicates unable to run analysis due to insufficient data. *Indicates Fisher’s exact test was used.

