## Supplemental Table 1 for "Examining the role of women’s engagement in khat production on child nutritional outcomes using longitudinal data in East Oromia, Ethiopia"

**S1 Table. Chi-square or Fisher’s exact test results for women’s engagement in khat production and women’s empowerment.**

| Variable 1 | Variable 2 | Chi-Square | P-Value |
| --- | --- | --- | --- |
| Baseline women’s engagement | Baseline women’s empowerment | ^ | ^ |
| Endline women’s engagement | Baseline women’s empowerment | 0.552 | 0.457 |
| Change in women’s engagement over time | Baseline women’s empowerment | ^ | ^ |
| Baseline women’s engagement | Endline women’s empowerment | 0.447 | 0.504 |
| Endline women’s engagement | Endline women’s empowerment | 1.637 | 0.201 |
| Change in women’s engagement over time | Endline women’s empowerment | ^ | 0.303^*^ |

^Indicates unable to run analysis due to insufficient data. ^*^Indicates Fisher’s exact test was used.
